# Associations of cholecystectomy with the risk of gastroesophageal reflux disease: a Mendelian randomization study

**DOI:** 10.1101/2024.03.17.24304416

**Authors:** Jin Qian, Huawei Xu, Jun Liu, Yihu Zheng

## Abstract

**Background:** Cholecystectomy is the standard surgery for patients with gallbladder disease, but the impact of cholecystectomy on gastroesophageal reflux (GERD) is not clear.

**Methods:** We obtained genetic variants associated with cholecystectomy at a genome-wide significant level (*P* value < 5 × 10^−8^) as instrumental variables (IVs) and performed Mendelian randomization (MR) to explore the relationship with GERD.

**Results:** The Inverse Variance Weighted analysis (IVW) showed that the risk of GERD in patients after cholecystectomy increased (OR = 2.19; 95% CI: 1.18 – 4.09). At the same time, the analysis results of weighted median (OR = 2.30; 95% CI: 1.51 – 3.48) and weighted mode (OR = 2.21; 95% CI: 1.42 – 3.45) were also consistent with the direction of the IVW analysis and were statistically significant (*P* < 0.05).

**Conclusions:** This study shows that patients who have undergone cholecystectomy are a susceptible population of GERD.

## Introduction

Cholecystectomy is the standard surgical method for a variety of gallbladder diseases, and is suitable for various acute and chronic cholecystitis, symptomatic gallbladder stones, gallbladder protuberant lesions, etc [1-3]. It is one of the most common surgeries in general surgery. Regardless of the region, the number of completed cholecystectomy surgeries completed each year is a very large number. Therefore, further research on the impact of cholecystectomy is particularly important. Unfortunately, most of the current research on the impact of cholecystectomy is limited to the short-term postoperative complications caused by surgical operations, and there are few studies on the non-surgical long-term complications caused by cholecystectomy. If there is a deeper understanding of this aspect, it will play a very important role in improving the postoperative diagnosis, treatment and follow-up of patients.

GERD is defined by recurrent and troublesome heartburn and regurgitation or GERD-specific complications and affects approximately 20% of the adult population in high-income countries [4]. Not only are the symptoms very troublesome and have a great impact on the quality of life of patients, long-term GERD also increases the risk of developing esophageal cancer [5].

Whether cholecystectomy is a risk factor for GERD has been eagerly discussed for decades. Previous clinical studies generally believed that cholecystectomy would not lead to an increase in GERD and would not constitute a risk of GERD [6, 7], but there were also reports that duodenogastric reflux in patients with cholelithiasis or previous cholecystectomy was more than that in the healthy control group [8].

MR analysis is a method that uses genetic variants strongly associated with exposure factors as IVs to infer the association between exposure factors and outcomes. Compared to traditional observational epidemiological studies, MR analysis is based on Mendelian inheritance laws, where parental alleles are randomly allocated to offspring. This can be regarded as a natural randomized controlled trial (RCT) that is less susceptible to confounding factors and has a higher level of evidence [9].

The purpose of this study is to investigate the potential link between cholecystectomy and GERD via MR analysis, so as to facilitate the further improvement and development of the diagnosis and treatment of patients requiring cholecystectomy in clinical practice.

## Methods

This study was reported in accordance with the guidelines of *Strengthening the reporting of observational studies in epidemiology using Mendelian randomization: the STROBE-MR statement* [10].

### Study design

The study of MR should be carried out based on three principal assumptions: (1) the relevance assumption: there is a strong correlation between the Instrumental variable (IV) and the exposure factor; (2) the exclusivity assumption: IVs were not associated with potential risk factors for outcomes; (3) the independence assumption: IVs influence results only through exposure. In this study, we used “cholecystectomy” as the exposure point for a comprehensive MR analysis and conducted sensitivity analyses to reveal the association between cholecystectomy and GERD. Three core hypotheses were appropriately addressed in our analysis. The research design framework is shown in the figure 1.

**Figure 1:**
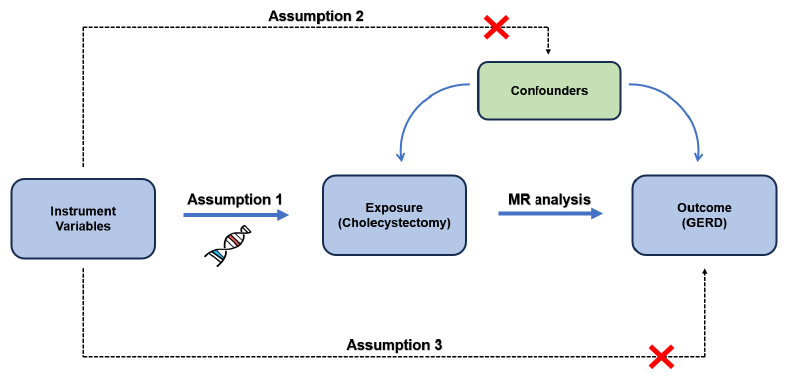
The basic principles of MR design

### Instrumental variable selections

In this study, we selected single nucleotide polymorphisms (SNPs) significantly associated with “cholecystectomy” as IV. We used a *P* < 5 × 10^−8^ cutoff to screen SNPs associated with the exposure, and evaluated linkage disequilibrium (LD) by calculating the r^2^ value. Further SNP screening was conducted based on LD and genomic regions (up/downstream: ± 10000kb, LD r^2^ < 0.001). In addition, we further evaluated the validity of each IV through the *F*-statistic, and the calculation formula is 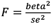, “beta” represents the magnitude of the impact of the IV on the exposure, and “se” is the corresponding standard error of “beta”. An *F*-statistic < 10 is considered as weak IV bias, and we eliminated SNPs with *F* < 10 to avoid the weak instruments bias [11].

### Statistical analysis and Sensitivity analysis

To explore whether there is a relationship between exposure and outcome, we used various methods such as random effects IVW, MR Egger, weighted median, and weighted mode for analysis and research. Among them, IVW is used as the main analysis method because it has recognized high statistical potency, stability, and can provide appropriate analysis in the presence of heterogeneity. The IVW method employs a meta-analysis approach, combining the Wald estimates of each SNP to obtain an overall estimate of the impact of exposure on the outcome [12]. IVW has two effect models, and the fixed-effect model or random-effect model is selected according to heterogeneity [13]. If significant heterogeneity (*P* < 0.05) is observed, the random-effect IVW model is used, otherwise, the fixed-effect model is used. Other methods such as MR Egger are used to supplement and verify the results of the IVW analysis. For example, the MR-Egger regression analysis can provide tests for unbalanced pleiotropy and heterogeneity. When there is pleiotropy, the MR-Egger estimate is more persuasive than the IVW estimate [14].

We used sensitivity analysis to test and correct the robustness of MR analysis. Heterogeneity was detected by Cochran’s Q test. The pleiotropy was evaluated using the intercept term derived from the MR-Egger regression. A leave-one-out analysis was performed to determine if the association estimate was driven by a single SNP.

All analyses were performed with the “Two Sample MR” package in R software.

### Data source

In this study, we selected the Genome-wide association study(GWAS)summary statistics with the largest sample size from the public website “open GWAS”(https://gwas.mrcieu.ac.uk/) developed by the MRC Integrative Epidemiology Unit (IEU) at the University of Bristol. The GWAS ID of cholecystectomy is “ukb-b-6235”, which includes 18,319 cases and 444,614 controls. The GWAS ID of GERD is “ebi-a-GCST90000514”, which includes 129,080 cases and 473,524 controls. All the GWAS selected in our study were conducted in the European population, and population stratification and other demographic variables (such as gender and age) were adjusted in the original GWAS analysis. Each GWAS has been approved by its respective ethical committee, and the data can be used without restriction.

## Result

Using cholecystectomy as the exposure and GERD as the outcome, we extracted 19 SNPs as IVs, and each IV was valid with an *F*-statistic > 10. As shown in table, the IVW analysis showed that the risk of GERD in patients after cholecystectomy increased (OR = 2.19; 95% CI: 1.18 - 4.09, *P* = 0.0132). At the same time, the analysis results of weighted median (OR = 2.30; 95% CI: 1.51 – 3.48, *P* = 9.15 × 10^−5^) and weighted mode (OR = 2.21; 95% CI: 1.42 – 3.45, *P* = 0.0024) were also consistent with the direction of the IVW analysis and were statistically significant (*P* < 0.05). The result of MR Egger analysis (OR = 2.40; 95% CI: 1.05 – 5.45, *P* = 0.0526) is consistent with the directions of the other analysis results, but it is not statistically significant.

In terms of sensitivity analysis, although heterogeneity was observed in the Cochran Q test for heterogeneity analysis (Q = 51.99, *P* = 2.06 × 10^−5^), the result of MR-Egger regression indicates that pleiotropy appears to be minimal (*P* = 0.743)(Table 2). Combined with Figure 2a, the Egger-intercept for MR-Egger was not statistically significantly different from 0, indicating that the presence of heterogeneity does not cause any pleiotropy bias in the MR estimate. However, through the leave-one-out analysis, we found that removing SNP rs11887534 would result in consistent but non-significant results, indicating that the association between them may be disrupted by this SNP, and the conclusion should be cautious (Figure 2b).

**Table 1.**
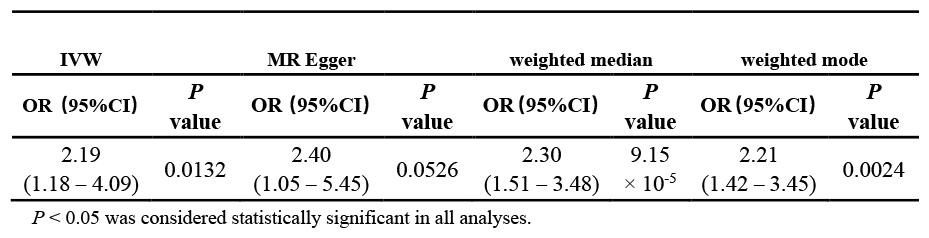
Result of MR analysis.

**Table 2.** Results of sensitivity analysis.

| Cochran Q test |  | MR-Egger regression |
| --- | --- | --- |
| Q | P value | P value |
| 51.99 | $2.06 \times 10^{-5}$ | 0.743 |

$P < 0.05$ was considered statistically significant in all analyses.

**Figure 2:**
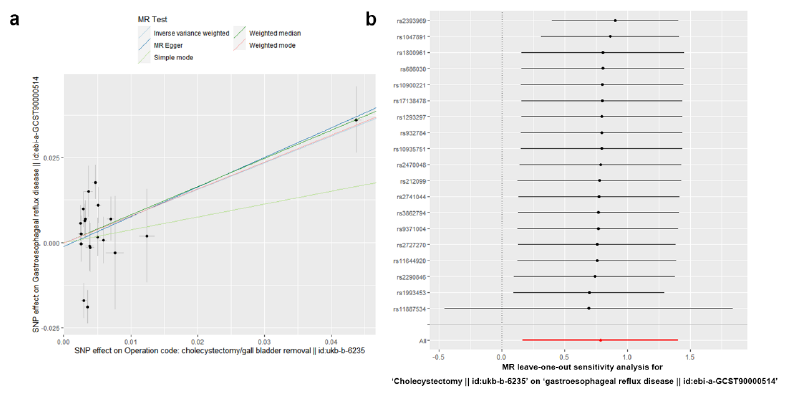
**(a)** The result of MR-Egger regression; **(b)** The result of leave-one-out analysis.

## Discussion

Despite the fact that cholecystectomy has now become a routine operation for the treatment of biliary tract diseases, which can effectively treat gallbladder diseases, it is still very important to further understand whether it will increase the risk of other diseases. This will affect the use of cholecystectomy by hepatobiliary surgeons, as well as the surgical options for some patients with gallbladder diseases, and has important clinical significance.

In the study of cholecystectomy and GERD, traditional studies in the past generally believed that there was no connection between cholecystectomy and GERD. However, in this study, we used MR analysis and large-scale database to analyze the data and came to a new conclusion that people who have undergone cholecystectomy are more likely to suffer from GERD than those who have not undergone cholecystectomy. Our contention is that the emergence of distinct conclusions as compared to those in the past can be attributed to the variance in research methodologies and the data being investigated. For example, in the study of cholecystectomy and GERD published by the Otto S. Lin team in the Annals of surgery in 2010, a reflux symptom questionnaire survey was adopted. Patients who underwent cholecystectomy filled in the reflux symptom score (RSS) and the gastrointestinal symptom rating scale (GSRS) 1 to 15 days before surgery and 4 to 12 weeks after surgery. Based on these data, the team concluded that cholecystectomy does not lead to an increase in reflux symptoms [6]. The research by Manifold, D.K.’s team in 2000 studied the connection between cholecystectomy and GERD by measuring the changes in gastric pH values at different time points, with the longest follow-up time only stopping at 3 months after the cholecystectomy [7]. It can be seen that most existing studies are limited to whether GERD occurs in the short term after cholecystectomy, and there is a lack of long-term follow-up data. It is still undetermined whether cholecystectomy will increase the risk of GERD occurring for a long time after the operation. In this study, we conducted the study by means of MR Analysis, which could not be affected by the postoperative follow-up time, so as to avoid the bias caused by the lack of postoperative follow-up time, which is a supplement and perfection of traditional clinical controlled studies.

MR analysis utilizes genes as IVs to more effectively control the influence of confounding factors. Traditional observational studies are often affected by reverse causation and selection bias, while MR analysis can more reliably infer the association between outcomes and exposures through the known association between genes and diseases. Moreover, as MR analysis is based on genetic variations in the population, it has higher external validity. In addition, compared to other interventional research methods, MR analysis typically does not involve individual interventions, thus having the characteristic of non-invasiveness. In this MR analysis, we utilized multiple analytical methods, including IVW, MR Egger, weighted median, and weighted mode, to conduct the examination. Almost all analyses show a consistent association, indicating that patients after cholecystectomy are more likely to suffer from GERD. In combination with traditional clinical controlled studies, this may occur over an extended period after the surgery. The possible reasons for this phenomenon are as follows: (1) bile reflux: The main function of the gallbladder is to store and release bile, which aids in the digestion of lipids. The removal of the gallbladder results in the direct flow of bile from the common bile duct into the duodenum, increasing the risk of bile reflux into the stomach [15]. Bile reflux into the stomach can stimulate the gastric mucosa, leading to increased acid secretion, thereby increasing the risk of GERD; (2) dysfunction of the antroduodenal motor unit: Cholecystectomy may cause dysfunction of the antroduodenal motor unit, making it easier for the contents of the duodenum to reflux back into the stomach and stimulate the gastric mucosa, which in turn leads to excessive acid secretion and increases the risk of GERD [16]; (3) gastric motility: After cholecystectomy, it may affect the neural regulation and motor function of the gastrointestinal tract, resulting in delayed gastric emptying or abnormal peristaltic movement of the esophagogastric junction, making it easier for gastric acid and gastric contents to reflux into the esophagus; (4) sphincter dysfunction: The lower esophageal sphincter and pyloric sphincter can prevent the reflux of gastric acid and gastric contents. After cholecystectomy, the function of these sphincters may be impaired, leading to improper functioning and an increased likelihood of GERD; (5) diet: After cholecystectomy, the patient’s ability to digest fats is reduced [17]. Consuming high-fat foods after surgery may be due to insufficient bile secretion, potentially causing indigestion and increasing the risk of GERD. It is worth noting that although the MR analysis shows an association between cholecystectomy and GERD, after excluding rs11887534, although the results are in the same direction, the results lose statistical significance, indicating that the association between cholecystectomy and GERD may be largely influenced by the rs11887534. In addition, all the subjects in this study are of European descent, and the conclusions of this study may not be applicable to other races.

To sum up, through this MR study, we believe that cholecystectomy increases the occurrence of GERD, and this phenomenon may occur for a relatively long time after the surgery. Although the MR analysis of cholecystectomy and GERD in this study presents a completely opposite result from traditional clinical controlled studies, we do not think this is a denial of the results of traditional clinical research. Rather, we see it as a complement to the previous research findings. This study fills the gap in the research on the possible occurrence of GERD conditions for a longer period after cholecystectomy in previous traditional clinical controlled trials through MR analysis, providing a further understanding of the relationship between cholecystectomy and GERD. Clinically, this finding is of great significance. It reminds doctors to consider the overall health status of patients and possible postoperative complications more cautiously when deciding whether to perform cholecystectomy. For patients who have undergone cholecystectomy, doctors may need to pay more attention to their gastrointestinal symptoms, especially symptoms related to GERD, in order to make timely diagnoses and treatments. In addition, this finding also provides new ideas for the treatment of GERD. Researchers can further explore the specific mechanism between cholecystectomy and GERD, develop targeted treatment methods or medications to improve the symptoms and quality of life of patients. This finding also reminds patients to pay attention to their own gastrointestinal health before and after undergoing cholecystectomy.

## Data Availability

All data produced are available online at

## Author contributions

Y.Z conceived and designed the current study., J.Q and J.L performed analyses. J.Q and W.X prepared the manuscript. J.Q and Y.Z reviewed and edited the manuscript. All authors read and approved the manuscript.

## Data availability

Publicly available summary statistics are obtained from https://gwas.mrcieu.ac.uk/.

## Acknowledgements

This research was conducted using data from Genome-wide association study(GWAS)summary statistics from the public website “open GWAS”, https://gwas.mrcieu.ac.uk/. We thank the participants, contributors, clinicians, and researchers for making data available for this study.

